# Phage-plasmid borne methionine tRNA ligase mediates epidemiologically relevant antimicrobial survival

**DOI:** 10.1101/2024.10.27.24316207

**Authors:** Yi Ling Tam, P. Malaka De Silva, Clare R. Barker, Ruizhe Li, Gherard Batisti Biffignandi, Leanne Santos, Charlotte E. Chong, Lewis C. E. Mason, Satheesh Nair, Paolo Ribeca, Sion C. Bayliss, Claire Jenkins, Somenath Bakshi, James P.J. Hall, Lauren Cowley, Kate S. Baker

## Abstract

Antimicrobial resistance (AMR) is a global public health crisis with few options for control. As such early identification of emerging bacterial strains capable of rapidly evolving AMR is key. Although antimicrobial tolerance and persistence are precursor phenotypes for AMR, little evidence exists to support their importance in real-world settings. Here we used bacterial genome wide association on national genomic surveillance data of the diarrhoeal, and World Health Organisation AMR priority pathogen, *Shigella sonnei* (n=3745) to agnostically identify common genetic signatures among lineages convergently evolving toward AMR (n=15). This revealed an association of an AMR trajectory with a multi-and highly variable second copy of *metG*, borne by a phage-plasmid we called pWPMR2. Further analyses revealed that pWPMR2 was present across clinically relevant enteric pathogens globally, including past and contemporary outbreaks, and that the additional-*metG* mechanism was present across multiple bacterial phyla. Functional microbiology, experimental evolution, and single-cell physiology studies confirmed that the expression of auxiliary *metG*, particularly the mutated version on pWPMR2, created a sub population of cells predisposed to survival in, and evolving resistance against, third generation cephalosporins (3GC). Thus, we demonstrate a novel mechanism of auxiliary *metG* carriage that predisposes bacteria to AMR with real world impacts. Furthermore, our approach is a timely example of using genomic epidemiology to rapidly guide functional microbiology studies in the era of routine genomic surveillance and also highlights several deficiencies in current AMR surveillance practices.

## Background

Antimicrobial resistance (AMR) is a global public health crisis, and microbial genomics has revealed that certain highly AMR bacterial lineages dominate epidemiologically. For example, *S. Typhimurium* Sequence Type 313 (ST313) in African invasive disease^1^, the pandemic *E. coli* ST131^2–5^, the carbapenemase associated *K. pneumoniae* ST258^6^, and Lineage III *S. sonnei* which has recently evolved extensive drug resistance (XDR)^7–9^. Once AMR has been acquired and disseminated in these successful clones, the resulting treatment complications make their control challenging. In an ideal world, we would use the imminent tsunami of genomic surveillance data to move toward predictive frameworks of AMR surveillance in which clones that are predisposed to AMR could be identified for timely intervention. However, this requires an understanding of the precursors or ‘stepping stones’ that occur before AMR evolves or is acquired.

Here, we leveraged an epidemiological scenario of convergent evolution of AMR among lineages of a highly clonal pathogen to agnostically identify pathogen-factors associated with an AMR trajectory (i.e. adapting to high levels of antimicrobial selection pressure). Specifically, the severe diarrhoeal pathogen, *Shigella*, has established over the last two decades as a sexually transmissible illness (STI) among men who have sex with men (MSM)^10^. This is particularly detectable in traditionally non-endemic, high-income countries, such as the United States and the United Kingdom, where an over-representation of closely related isolates derived from males without a recent history of travel is used to identify lineages that are circulating in sexual transmission networks (STN)^11,12^. These STN lineages comprise frequently co-circulating polyphyletic lineages in an otherwise geographically associated population structure where isolates from diverse endemic settings (sampled from returning travellers) are phylogenetically proximate^9,10,13,14^. The antimicrobial use in STN is evidenced empirically (one report revealed 43% of shigellosis-affected MSM had received antimicrobials in the last 6 months^15^) and the convergent acquisition of AMR plasmids is frequently bystander resistance resulting from high intensity treatment for other traditional STIs (e.g. gonorrhoea, chlamydia)^9,10,16–19^. Concerningly, it is also clear that these lineages spread globally over short time frames and share plasmids among themselves and with other pathogen groups. Thus, convergently evolving genetic traits among co-circulating STN Lineages of *Shigella* represent potential adaptations to high selective pressure from antimicrobial use with a high potential for global spread.

In this study, we identify convergently evolving factors across 15 STN lineages among some 3,745 *Shigella sonnei* isolates from routine surveillance from 2004 to 2020 in the United Kingdom using bacterial Genome Wide Association Study (bGWAS) to identify ‘stepping stones’ to AMR. We identified variant copies of methionine tRNA ligase (*metG*) carried on an SSU5-like phage-plasmid, which we called pWPMR2, as associated with STN Lineages (a proxy for high antimicrobial selection). Further *in silico* analyses showed that both pWPMR2 and other plasmid-borne *metG* are spread globally in other enteric bacteria and bacterial phyla. We demonstrate how auxiliary *metG* expression and variation create bacteria with survivor phenotypes that predispose bacteria to evolve AMR against third generation cephalosporins.

### Phage-plasmid borne *metG* is associated with an AMR trajectory

To identify genetic features associated with AMR emergence, we first identified convergently evolving STN lineages of *S. sonnei*. To do this, we constructed a recombination-free maximum-likelihood phylogenetic tree of S*. sonnei* isolates (n=3,745) from routine surveillance in the United Kingdom between 2004 and 2020 (Fig. 1a, Supplementary Table 1). Adapting validated approaches, we then used demographic data on patients’ age, sex, and travel history for ancestral state reconstruction to identify 15 lineages circulating in STN (hereafter STN lineages, Fig. 1a, Table1, Extended Data Fig. 4, Methods). We identified 15 STN lineages of variable sizes which, consistent with the increased antimicrobial selection pressure in STN, had a significantly higher proportion of isolates containing genes conferring AMR against (any of) the three main clinically relevant antimicrobials for *Shigella*^20^; ciprofloxacin (CIP), azithromycin (AZM), and ceftriaxone (CRO) (74% [n=765/1,028] compared with 37% [n=1011/2,717], *P* <0.001, Table 1, Supplementary Table 1). Alongside the distribution of these resistances in five, six, and five of the 15 STN lineages respectively, this confirms that *Shigella* circulating in STN are convergently evolving toward AMR, consistent with previous studies^10,17,18,21–23^.

**Fig. 1.**
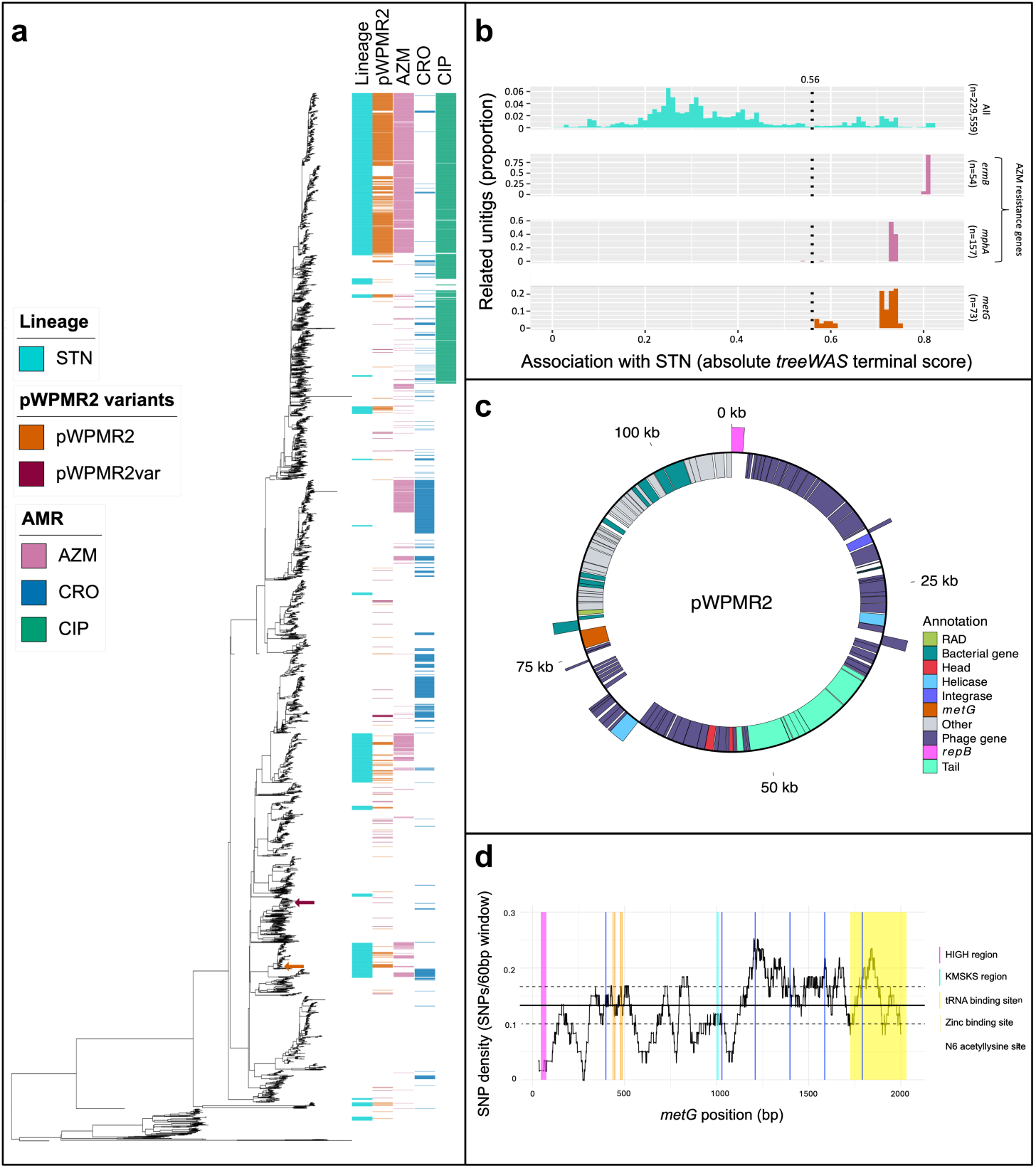
Variable metG borne on pWPMR2 is associated with bacterial lineages on an AMR trajectory. **a,** A midpoint-rooted maximum likelihood phylogenetic tree constructed from 32,294 recombination-free polymorphic sites shows the evolutionary relationships among 3,745 routine genomic surveillance *S. sonnei* isolates. The adjacent metadata columns show (from left to right): STN lineages (i.e. circulating in sexual transmission networks); the presence of pWPMR2 or pWPMR2var, and the presence of genotypically predicted resistance against three key antimicrobial treatments; azithromycin (AZM), ceftriaxone (CRO), and ciprofloxacin (CIP). The two isolates used to nanopore sequence pWPMR2 and pWPMR2var are indicated by arrows. **b,** Frequency histograms show the proportional density of unitgs with different Genome Wide Association Scores (absolute *treeWAS* terminal score). Distributions are shown for all unitigs with ≥0.05 minor allele frequency among the isolates, and unitigs relating to the AZM resistance genes *mphA* and *ermB* (acting here as positive controls) and *metG*. Unitigs with association to STN lineages were defined as those with an association score higher than 0.56 as indicated by the vertical dotted line. **c,** A genetic map of the phage-plasmid pWPMR2 showing forward (outer) and reverse (inner) open reading frames coloured by function according to the inlaid key. **d,** The variation in *metG* across the length of the gene among 650 pWPMR2/pWPMR2var-containing isolates. Median (solid) and interquartile-range quantiles (dashed) of SNP-density across the gene are indicated by horizontal lines and a sliding 60bp window of SNP-density (undulating line).

**Table 1.**
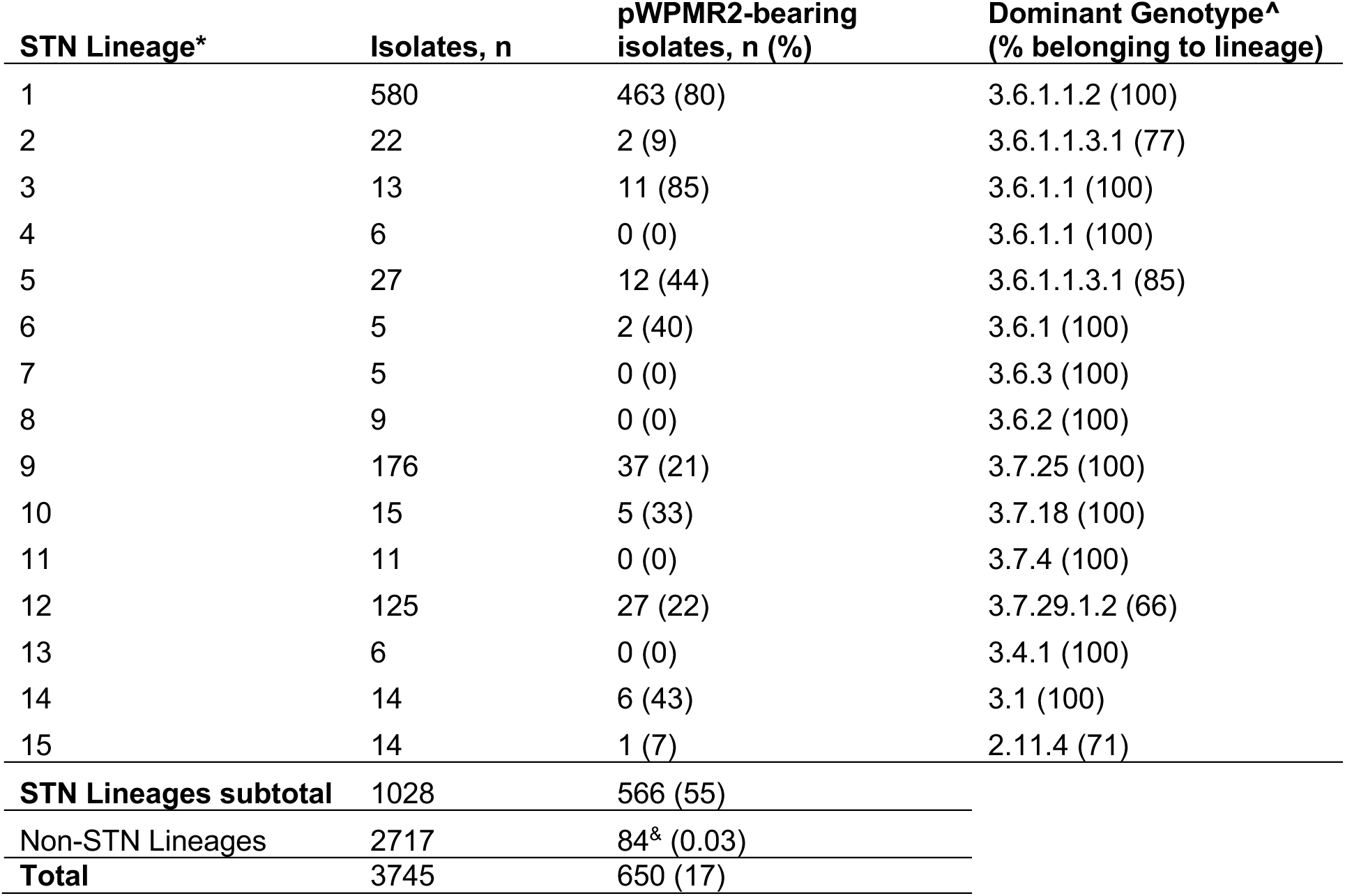
Characteristics of *S. sonnei* lineages circulating in STNs. * Arbitrarily named from top to bottom of tree in Fig. 1a – see Supplementary Data for individual assignments ^ Genotyping performed according to ^31,32^ ^&^ 53 of which were pWPMR2var

We then quantified the association of unique genetic sequences (i.e. unitigs from a compiled assembly graph of the 3,745 isolates) with membership of STN Lineages using *treeWAS*^24^. There were 229,559 unitigs with a minor allele frequency filter of 0.05 and had a distribution whereby the majority of unitigs (84%, n=192,832) had an absolute terminal association score of <0.56 (Fig. 1b, Supplementary Tables 2 and 3A, similar results with ‘subsequent score’ Extended Data Fig. 1). We then considered unitigs with scores over this threshold (n=36,727) as ‘highly associated’ with STN and explored their annotations. To do so, we mapped highly associated unitigs (n=36,727) to a concatenated reference containing the bacterial chromosome and the known AMR-plasmid pKSR100^17^ and retrieved the annotations of mapping locations. Overall, 544 highly associated unitigs mapped to 50 chromosomal genes and 12 intergenic regions (Extended Data Fig. 2). The gene with the strongest support (i.e. most unitigs with the highest terminal score, Extended Data Fig. 2, Supplementary Data 3) was *metG.* This gene shared similar association scores with the AZM resistance genes *mphA* and *ermB* (Fig. 1b) which act as positive controls as they are known to have emerged globally on pKSR100 among *Shigella* in STN (Fig. 1a)^10,17,18,21–23^. The gene *metG* encodes methionine tRNA ligase, which plays a core role in translation, linking the amino acid methionine (a start codon amino acid) to its cognate tRNA molecule (and is thus also involved in chain elongation). In laboratory settings, disruption to *metG* alters growth characteristics and is considered a canonical mutational route to antimicrobial tolerance (i.e. short-term survival against killing concentrations of antimicrobials without a shift in AMR phenotype^25–28^, which enables subsequent evolution of AMR^29^). For these reasons, we hypothesised that the auxiliary *metG* was assisting *Shigella* lineages during circulation in the high antimicrobial selection environment of STNs.

Initial bioinformatic analyses of coverage depth and variation in *metG* revealed both copy number and Single Nucleotide Polymorphisms (SNPs) variation (Extended Data Figs 5 & 6) which were challenging to interrogate with short-read data. Hence, we conducted complementary long-read sequencing of STN Lineage 12 strain SRR5005407 (Fig. 1a) which revealed that the extra copies of *metG* were carried on a 108,107bp circular sequence which we called pWPMR2 (GenBank accession: PQ180110). The pWPMR2 sequence was found to contain both plasmid replication and phage structural genes (i.e. capsid, tail) indicating a phage-plasmid (Fig. 1c). Furthermore, pWPMR2, also contained a viral anti-defence system (RAD^30^) consistent with a phage lifestyle. Mapping of our data to the new SRR5005407 genome reference confirmed that many highly associated unitigs (36%, n=13,187/36,727) belonged to pWPMR2 (Extended Data Fig. 3). Mapping coverage across our dataset revealed variation in pWPMR2 copy number and coverage (Extended Data Fig. 5, Supplementary Table 1). We inferred that 650 *S. sonnei* isolates contained pWPMR2 (using frequency distribution-informed cut offs, see Extended Data Fig. 5) which mostly (87%, n=566/650, *P* <2.2e-16) belonged to STN lineages (Fig. 1a, Table 1). Furthermore, while most pWPMR2-containing isolates contained one to two copies of pWPMR2, there was a wide range of relative depth (Extended Data Fig. 5b), suggesting possible instability of pWPMR2 carriage. Although this could also potentially arise from a laboratory artefact, pWPMR2 instability would be consistent with previous studies showing that acquisition of essential genes can compromise plasmid inheritance^31^. The generalisation of *metG* encoding on pWPMR2 across isolates was confirmed by linear regression of the mapping depth of *metG* and pWPMR2 (R^2^ = 0.91, *P* <2.2e-16, Extended Data Fig. 5c). These analyses also suggested the presence of a *metG*-bearing pWPMR2 variant (pWPMR2var, GenBank accession: PQ521219 from isolate SRR10313630, Extended Data Fig. 8) with reduced (∼0.75) coverage that was mostly found outside of STN (Fig. 1a, Table 1, Supplementary Table 1). No known AMR or virulence genes were present on pWPMR2, so we continued with our GWAS-guided hypothesis that auxiliary *metG* expression was driving a phenotypic shift relevant for adaptation to antimicrobial selection pressure.

An analysis of SNPs across *metG* among the isolates revealed that the auxiliary *metG* on pWPMR2 exhibited significant genetic variation relative to the essential chromosomal version. Specifically, isolates without pWPMR2 (i.e. only chromosomal *metG*) showed high levels of conservation, with only 12 of 3095 isolates containing SNPs across five sites in *metG*; four of which introduced non-synonymous changes (found in non-STN Lineages not phylogenetically proximate to STN Lineages), and one synonymous change in seven isolates of STN Lineage 15. Contrastingly, isolates that contained pWPMR2 had significant numbers of SNPs (0-118 per isolate, mean 66) affecting a total of 250 sites across the 2034bp gene, at minor allele frequencies consistent with a low-copy number mobile genetic element, with SNPs generally being phylogenetically grouped (Extended Data Fig. 6). A sliding window analysis of the variation (density of SNPs) on *metG* showed relative sequence conservation in certain key functional areas of the gene (such as the HIGH and KMSK amino acid motifs) and comparatively higher variation in the N6-acetyllysine sites (Fig. 1d), which might alter *metG* post-translational modification and abundance^32,33^, with a knock-on effect on overall protein production. While quantitating gene variation in multicopy genes using short read data is complex, our complete genome sequencing confirmed that the *metG* borne by pWPMR2 in our reference isolate SRR5005407 (see *metG*_PP_ below) contained 115 SNPs, 6 of which were non-synonymous. This highlights that the additional *metG* on pWPMR2 experiences significant genetic variation relative to single chromosomal copies.

The origins of this increased genetic variation are unknown but may result from multiple drivers e.g. increased replication/transmission of pWPMR2, increased mutation rates. One potential source of variation is the possibility of multiple introductions of *metG* from an external reservoir, which is supported by the presence of pWPMR2 and pWPMR2var. To examine the possibility of multiple introductions, we phylogenetically reconstructed the evolutionary relationships among *metG* located on pWPMR2/pWPMR2var in our surveillance dataset, alongside those on plasmids of various bacterial species in a public plasmid database (PLSDB, more below). This revealed that while there was evidence of multiple introductions of *metG* into *S. sonnei*, it is likely that a single introduction, associated with pWPMR2, made into *S. sonnei* circulating among sexual transmission networks which was followed by clonal expansion (Extended Data Fig. 7, Supplementary Table 4). This analysis did, however, suggest multiple potential introductions of pWPMR2var, highlighting the need for further extensive study in *metG*-bearing mobile genetic elements (more below).

### pWPMR2 and auxiliary metG are found globally in other clinically relevant organisms

Having associated a novel *metG*-bearing mobile genetic element with genomic lineages on an AMR trajectory, we then explored the extent to which pWPMR2 may have spread among clinically relevant organisms. A comparison of near-identical relatives of pWPMR2 in public databases revealed that it was present in multiple other major outbreaks of *Shigella* and Shiga-toxigenic *E. coli* and may have links with *E. albertii.* Specifically, it was near-identical (>99% similarity) to several undescribed plasmids from MSM-associated outbreaks of *S. sonnei*^33^ and O117:H7 Shiga toxin-producing *E. coli* (STEC)^34^, across a broad geographical range (Fig. 2a). A closely related element was also found to be present in an isolate belonging to an STEC O114:H4 outbreak in Georgia in 2009^35^, as well as an AZM-resistance bearing commensal *E. coli* strain (ST62) from a healthy college student in the USA^36^. Outside of *Shigella* and *E. coli* the closest relatives were found in *E. albertii* where multiple regions of pWPMR2 were absent (Extended Data Fig. 9). This demonstrated that pWPMR2 is a widely distributed mobile genetic element associated with important outbreaks of clinical disease as well as commensal organisms.

**Fig. 2.**
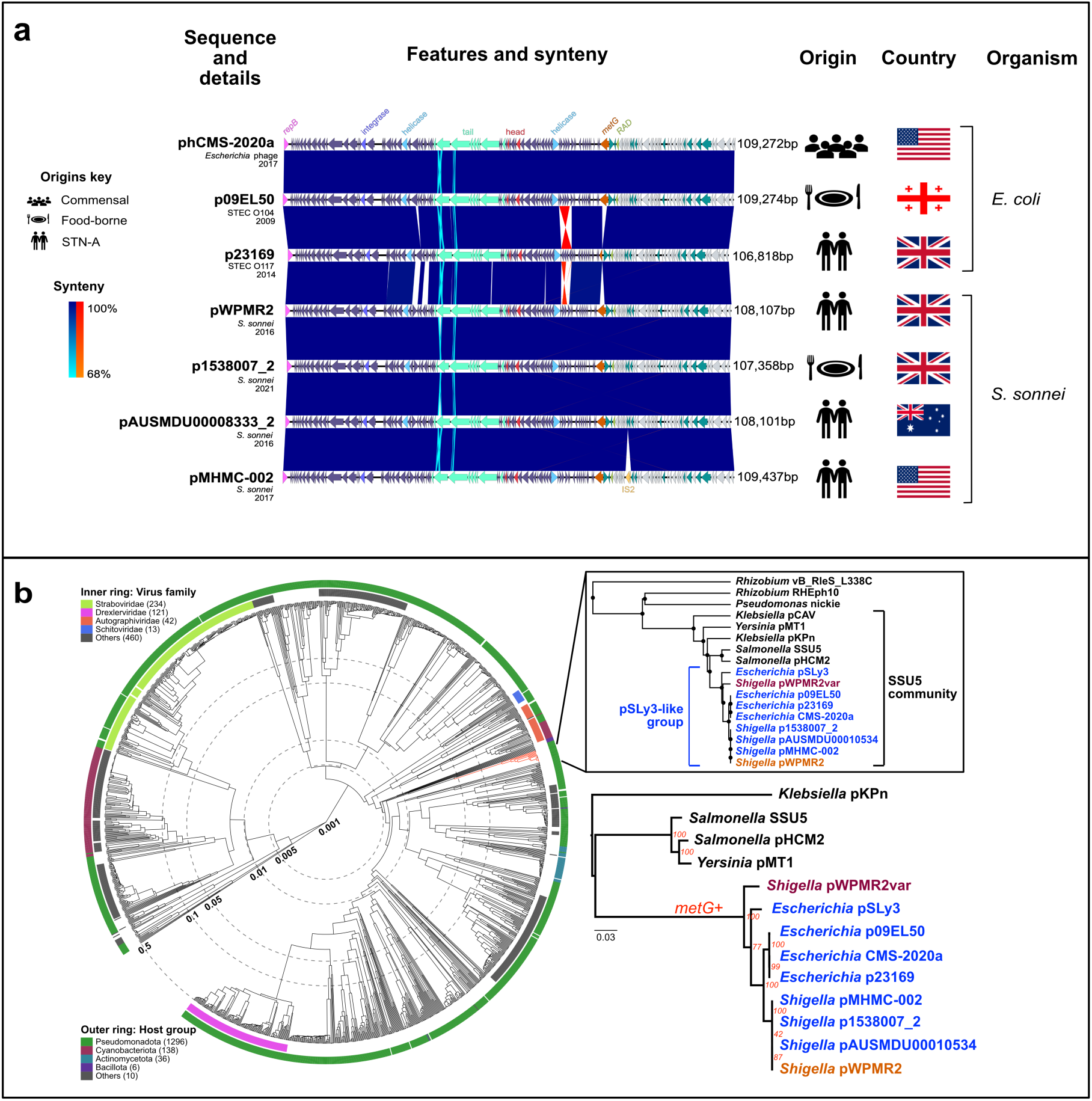
Epidemiological and taxonomic relatives of pWPMR2. **a,** Comparison of pWPMR2 with its most similar publicly available sequences (labelled leftmost) and their epidemiological details. The sequence comparison is shown as linearised replicons starting with *repB* gene. Certain relevant gene features are shown in colour and labelled uppermost. Colour blocks intervening between sequences showing colinear (blue scale) and inverted (red scale) synteny according to the inlaid coloured gradient keys. The origin of the sequence, country and organism are shown to the right of sequences. The symbol key for origin is shown leftmost. Other available details for the sequences (e.g. subtype, year of isolation) are shown under the sequence label. STEC = Shiga toxigenic *Escherichia coli*. STN-A = Sexual Transmission Network associated. **b,** The circular cladogram shows comparison among 1,512 related reference viral sequences (see methods). The branch lengths show genetic relatedness based on tBLASTx comparison (embedded in ViPTree) on a non-linear scale. The inner and outer rings show virus family and host group according to the inlaid keys, and the SSU5 community is highlighted as a red branch. The pop out shows a zoomed-in view of the clade containing SSU5 and its nearest relatives, highlighting the pSLy3 group (blue), containing pWPMR2 (orange) and pWPMR2var (claret). The lowest dendrogram is a maximum-likelihood phylogenetic tree of the SSU5 community, with bootstrap support values and *metG* acquisition indicated in red.

To further explore its association with clinically relevant organisms, we determined the taxonomy of pWPMR2 and screened all genomes of enteric infections in the UK isolated between 2016 and 2021 (n=66,929) for evidence of pWPMR2-like phage plasmids. Taxonomically, pWPMR2 was found to belong to the SSU5 super-community of phage-plasmids that are associated with species of the *Enterobacteriaceae* family^37^. Comparison of its genome-wide sequence similarity with other phage genomes showed that pWPMR2 belonged within the pSLy3 group (Fig. 2b, Supplementary Table 5). The screen of clinical isolates from enteric infections revealed that while SSU5 phage-plasmids were present in isolates from all genera, the pSLy3-like group was primarily associated with *S. sonnei* (18% of isolates), *E. albertii* (8%), *E. coli* (2%), and other *Shigella* species (0.5%, Table 2). This indicates the mobilisation of pWPMR2 across multiple enteric pathogen species, consistent with previous observations for AMR plasmids^17,38^.

**Table 2.** The presence of SSU5-like phage plasmid subtypes across enteric bacteria in the UK. ^1^ restricted to *Klebsiella* in GenBank ^2^ restricted to *Yersinia* in GenBank.

| Species | Genomes (n) | SSU5 supercommunity phage plasmid group containing (n) |  |  |  |  |  |
| --- | --- | --- | --- | --- | --- | --- | --- |
|  |  | pHCM2 | pSLy3 | pKpn<sup>1</sup> | pCAV<sup>1</sup> | pMT1<sup>2</sup> | Other |
| *S. enterica* | 47,495 | 539 | 0 | 0 | 0 | 0 | 2 |
| *E. coli* | 11,990 | 5 | 236 | 0 | 0 | 0 | 0 |
| *E. albertii* | 173 | 0 | 8 | 0 | 0 | 0 | 0 |
| *S. sonnei* | 3,427 | 2 | 628 | 0 | 0 | 0 | 0 |
| *S. flexneri* | 3,259 | 1 | 15 | 0 | 0 | 0 | 0 |
| *S. dysenteriae* | 196 | 0 | 0 | 0 | 0 | 0 | 0 |
| *S. boydii* | 289 | 0 | 4 | 0 | 0 | 0 | 0 |
| **Total** | **66,829** | **547** | **891** | **0** | **0** | **0** | **2** |

Given this evidence that pWPMR2 could spread across pathogens and our hypothesis that it was predisposing *Shigella* to AMR, we more fully characterised the temporospatial spread of pWPMR2. We first determined the temporal relationship of pWPMR2 acquisition with AMR across our 3,745 *S. sonnei* surveillance isolates (Fig. 3a). This showed that pWPMR2 entered the *S. sonnei* population earlier than, and then had similar temporal increases to, key AMR profiles (e.g. the plasmid-borne AZM and CRO resistances, and CIP resistance). Furthermore, in a Genotype specific analysis we found that among the 20 (of 67 total) *S. sonnei* Genotypes containing pWPMR2 and an AMR determinant (i.e. against AZM, CRO, CIP) these were more often (n= 17/26) Genotypes where pWPMR2 acquisition preceded AMR acquisition in the than AMR acquisition preceding pWPMR2 acquisition (n=6/26, with 3/26 Genotypes acquiring both pWPMR2 and AMR in the same year, Fig. 3b). The stepwise acquisition was particularly clear in the internationally disseminated XDR Genotype 3.6.1.1.2 (see STN Lineage 1 in Fig. 1a, Supplementary Table 1^38^). This pre-AMR acquisition is consistent with pWPMR2 conferring a phenotype relevant to the emergence of AMR. To explore spatial spread, we screened for pWPMR2 in over 2 million bacterial genome sequences in the public domain (Methods), and found it was spread among some 2,028 *E. coli* and *Shigella* isolates from broad geographical areas, often in isolates derived from public health surveillance (Fig. 3c, Extended Data Fig. 10, Supplementary Table 6). This built further genomic epidemiological evidence that pWPMR2 is an emerging mobile genetic element capable of spreading among pathogenic organisms.

**Fig. 3.**
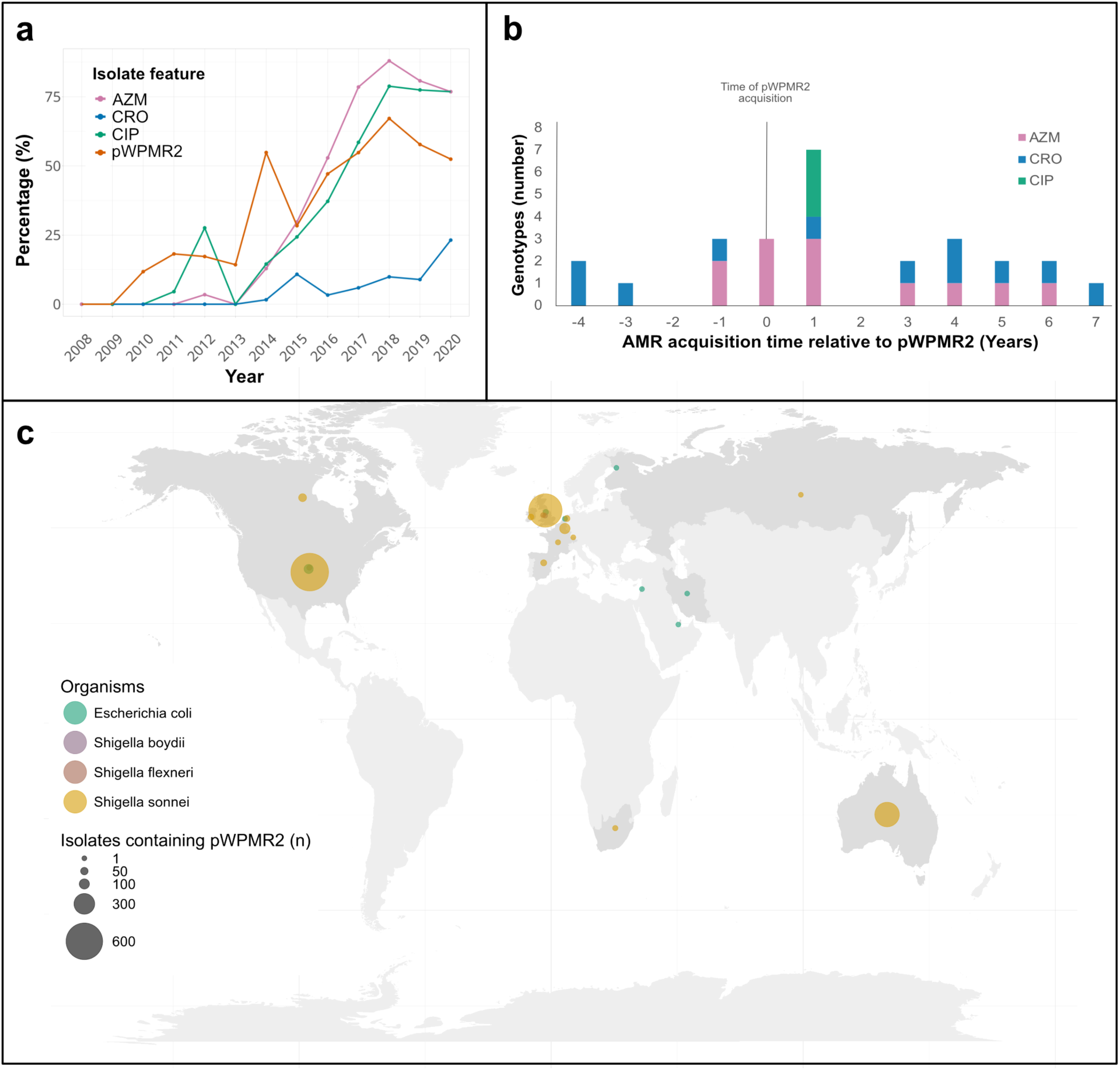
Temporal and spatial patterns of pWPMR2. **a,** The growth in the percentage of isolates containing pWPMR2, and resistance markers against three key antimicrobials among 3,745 *S. sonnei* surveillance isolates is shown over time according to the inlaid key, highlighting the early ingress of pWPMR2 into the *S. sonnei* population. **b,** The temporal relationships of pWPMR2 and AMR acquisition within individual Genotypes of *S. sonnei.* The frequency histogram shows the time in years between acquisition of pWPMR2 and acquisition of AMR determinants, coloured according to the inlaid key, among 20 *S. sonnei* Genotypes that contained both pWPMR2 and AMR. **c,** The global distribution of 2,028 pWPMR2-containing isolates in public databases is shown with countries containing sequences highlighted in darker grey. Bubbles over country centroids are shown scaled by the number of isolates and coloured by the organism of origin according to the inlaid keys.

To explore the relevance of auxiliary *metG* in the broader bacterial context, we screened sequences (n=59,895) from the PLSDB plasmid database^39^ for *metG*, and other tRNA ligases. This revealed that tRNA ligase homologues were found at low levels on plasmids (n=513/59895, 0.86%, Supplementary Table 7). Following filtration for potential chromosomal contamination (Methods), five tRNA ligases were found on an appreciable number (≥20) of plasmids: *alaS*, *cysS*, *metG*, *serS*, *thrS*. Of these, *metG* was outstanding as the most frequently observed tRNA ligase, detected more often than all the other tRNA ligases put together, and the only one found >5 times in >1 Family. Specifically, *metG* encoding plasmids were predominantly found in Enterobacteriaceae (n=172/29242, 0.59%) and Burkholderiaceae (*Ralstonia* spp., n=95/652, 14.57%). While the Enterobacteriaceae plasmids were overwhelmingly (n=165/172) recorded in the database as Incompatibility group F, B replicon type (FIB) phage-plasmids (consistent with pWPMR2 detection), the *Ralstonia* plasmids were putative chromids (large, domesticated plasmids associated with environmental adaptation) ranging from 1.78 Mb to 2.2 Mb (^40^, Extended Data Fig. 11), and their *metG* sequences were (generally) phylogenetically distinct from those of *Enterobacteriaceae* (Extended Data Figure 7). As widespread plant pathogens however, *Ralstonia* can experience various antimicrobial stresses, including molecules naturally secreted from competing soil microorganisms, chemical agricultural inputs, and environmental heavy metals, so they may also be served by contingent functional phenotypes conferred by auxiliary *metG* that could enhance antimicrobial tolerance^41^.

In contrast, when this screening was repeated for databases containing bacteriophage genomes, *metG* was not detected in any genome other than the pWPMR2-like *Escherichia* phage CMS-2020a (Fig. 2a, though some other tRNA ligases were more common [Supplementary Table 8]). While phage-plasmids lacked representation in such curated sequence databases^37,42,43^, a recent large-scale computational identification of phage-plasmids and analysis of their pangenomes found *metG* present only in the pSLy3 subgroup of the SSU5 supercommunity^37^, though other tRNA ligases are present in different phage-plasmid groups^37,44^. This was reinforced by our screening of UK clinical enteric bacteria where of the 1440 detected SSU5-like phage-plasmids (Table 2), only the members of the pSLy3 subgroup carried a copy of *metG*. Therefore, *metG* appears not to be a widespread feature among either bacteriophages or phage-plasmids.

### Mutated *metG* confers advantages in the presence of relevant antimicrobials

Having determined that *metG* was borne on multiple mobile genetic elements, we sought to explore phenotypes associated with auxiliary *metG* expression and the observed genetic variation in *metG* on pWPMR2. Specifically, we conducted laboratory experiments on growth characteristics and antimicrobial response phenotypes of *E*. *coli* SB8 (a derivative of MG1655, see methods) strains containing inducible vectors that expressed wildtype (i.e. *S*. *sonnei* chromosomal) *metG* (hereafter *metG*_WT_) and the mutated version found on pWPMR2 (hereafter *metG*_PP_). We first confirmed that *metG* expression did not result in a change in AMR phenotypes, finding Minimum Inhibitory Concentrations (MIC) consistent across strains (Extended Data Table 1). Bulk growth experiments then revealed that induction of either *metG*_WT_ or *metG*_PP_ reduced overall bacterial growth, primarily through both *metG*_WT_ and *metG*_PP_ introducing an increased lag time, consistent with a previously reported tolerance by lag phenotype^45^ (Fig. 4a). Using these constructs, we then determined the impact of auxiliary *metG* expression (by comparing *metG*_WT_ with expression induced and uninduced) and *metG* variation (by comparing induced *metG*_WT_ with induced *metG*_PP_) in various phenotypic assays.

**Fig. 4.**
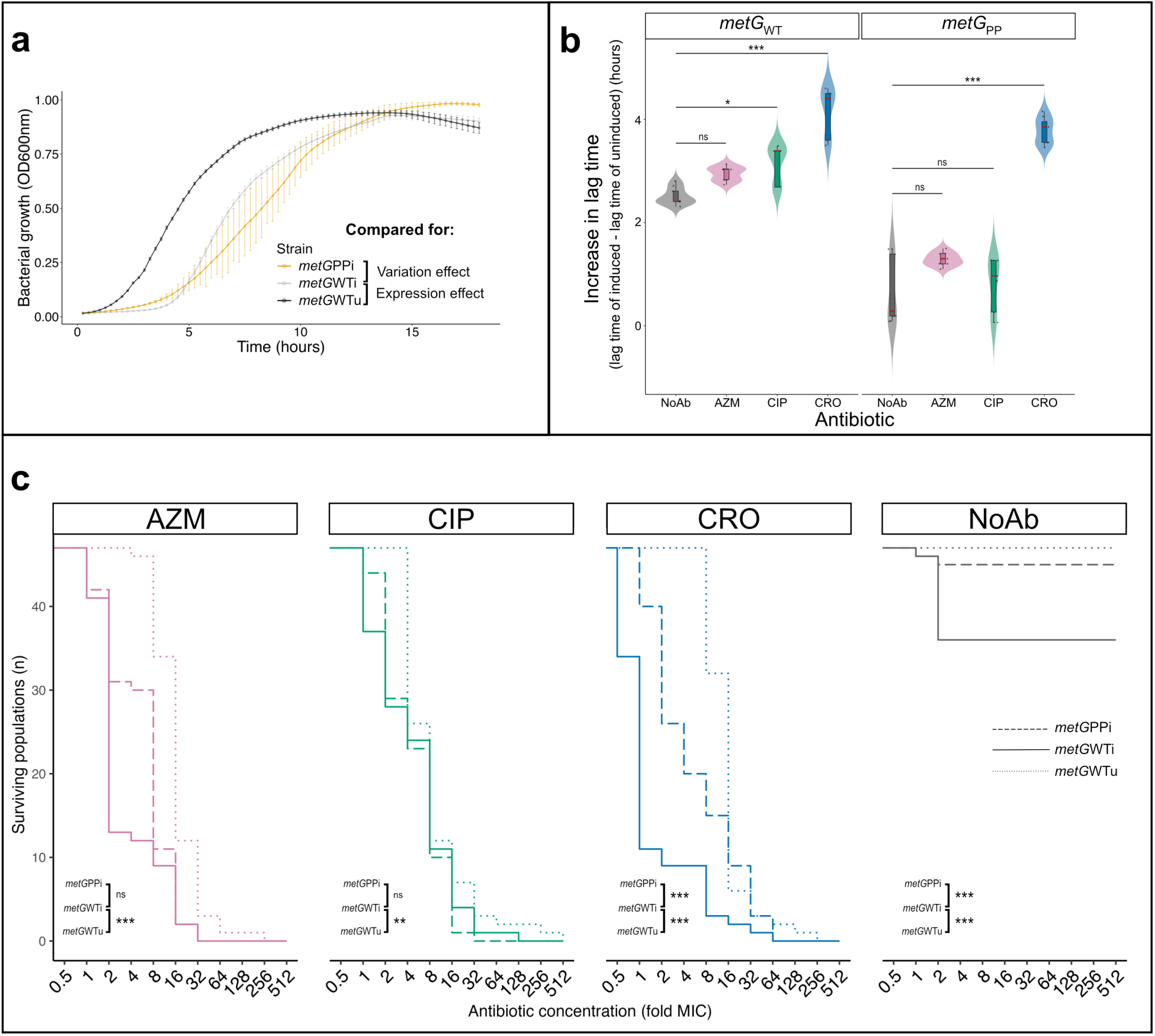
Auxiliary *metG* expression and variation generates altered growth profiles and antimicrobial response phenotypes. **a,** Growth over time for *E. coli* SB8 constructs with auxiliary *metG* _WT_ expression induced (*metG*WTi) and the respective (i.e. uninduced) control (metGWTu) along with the induced pWPMR2 phage-plasmid version of *metG* (*metG*PPi). The error bars show the 95% confidence limits of the mean (marker) of 9 data points (see methods) **b,** Increase in lag time of *E. coli* SB8 strains expressing *metG* constructs (WT and PP) in the presence of subinhibitory concentrations of key treatment antimicrobials (0.5x MIC) compared to the respective uninduced versions. The leftmost plot shows the increase in lag time in *metG*_WT_ bearing strains in the presence of sub-MIC concentrations of the indicated antimicrobials while the rightmost plot shows the same for the *metG*_PP_ bearing strains. Box and whisker plots within the violin plots show the median (centre line in red) with the whiskers indicating the 25^th^ and 75^th^ percentile values. Individual data points are also shown within the violin plots. Asterisks denote the statistical significance from pairwise comparisons calculated using Kruskal-Wallis test and corrected by Dunn’s test (Benjamini-Hochberg correction) where * denotes *P* <0.05, *** denotes *P* <0.001 and “ns” denotes no statistical significance. **c,** Population survival curves of *E. coli* SB8 carrying plasmid constructs of *metG*_WT_ uninduced (dotted), *metG*_WT_ induced (solid) and *metG*_PP_ (dashed) in the presence of indicated antimicrobials starting at 0.5x MIC and going up to 512x MIC. The asterisks for the relevant comparisons are shown in the sub panels where ** denotes *P <*0.01 and *** denotes *P* <0.001 and ns denotes no statistical significance as determined by log-rank test. Full statistical analyses including Hazard Ratios are included in Supplementary Table 9. CRO = ceftriaxone, AZM = azithromycin, CIP = ciprofloxacin

Although MICs had not changed, we hypothesised that alternative antimicrobial phenotypes might be particularly relevant for evolution in surviving in STN given the high levels of antimicrobial use as well as long half-life (e.g. 68 hours for azithromycin^46^), high plasma concentrations (e.g. ceftriaxone 13-239µg/ml after first dose^47^) and widespread practice of bolus dosing for STIs^48,49^. To explore this possibility, we first explored a potential benefit of auxiliary *metG* in the presence of sub-inhibitory concentrations of antimicrobials (0.5x MIC). Consistent with phenotypes previously associated with *metG* mutations, we saw an increase in lag time with the expression of *metG* (Fig. 4b). Expression of *metG*_WT_ in the absence of antimicrobials extended lag time by more than two hours, and sub-MIC CIP and CRO extended this further (*P* = 0.0370 and *P* ≤0.001 respectively, Kruskal-Wallis tests, Dunn’s adjusted). Contrastingly, *metG*_PP_ had lower lag time penalties compared to *metG*_WT_, offered increased variance in the no antimicrobial and CIP conditions, and selectively increased lag in the presence of CRO (*P* ≤0.001, Kruskal-Wallis test, Dunn’s adjusted, Fig. 4b). Collectively, these results suggest that *metG_PP_* might offer a tolerance by lag phenotype specific to CRO and an increased variance in population lag times which might offer survival advantages under antimicrobial exposure.

We then determined the role of *metG* expression and variation in the adaptation to supra-MIC concentrations by conducting experimental evolution of 47 independent populations of *E. coli* expressing *metG*_WT_ and *metG*_PP_ for eleven days, starting at 0.5x MIC and doubling daily to 512x MIC. None of the populations survived until the end for any antimicrobial condition (Fig. 4c) and expression of either *metG* had an adverse effect on the number of surviving populations (Fig. 4c, Extended Data Fig. 12). However, we also saw a statistically significant increase in the number of surviving populations as a result of the variation found on *metG*_PP_ (relative to *metG*_WT_) in the presence of CRO (Fig. 4c). This is consistent with the results above suggesting a potentially ‘special’ effect for pWPMR2 in tolerating CRO. In fact, when we truncated our experimental data to clinically relevant concentrations of CRO^47^ (≤ 16x MIC), we found no statistically significant survival difference among the induced and uninduced populations (Fig. 4c, Extended Data Fig. 13). This further highlighted the important role that *metG*_PP_ could be playing in bacterial populations adapting to CRO exposure and motivated us to investigate the effect at single cell resolution.

### *metG*_PP_ facilitated survival in CRO is mediated by multiple phenotypes

To further resolve the observed advantages in response to CRO, we conducted time-resolved single-cell analysis of the CRO response in strains carrying inducible *metG*_WT_ or *metG*_PP_ plasmids. Briefly, plasmids were introduced into *E. coli* strains carrying a constitutively expressed (under the PrpsL promoter), chromosomally integrated Cyan or Yellow Fluorescence Protein^50^ to distinguish pooled strains, and a plasmid-free background strain, constitutively expressing Red Fluorescence Protein (Fig. 5a). The mixed population of three strains was loaded into two separate lanes of a high-throughput microfluidic device^51^ such that cells were loaded into trenches randomly within each lane to eliminate systematic biases (Extended Data Fig. 14). In this setup, cells in one lane were exposed to media containing the *metG* inducer, while cells in the other lane received media without the inducer. Notably, in the absence of antimicrobials, there was no impact on cell growth upon induction of *metG*_PP_ (Extended Data Fig. 15). The narrow trench design of the microfluidic device ensured that the progeny of the cell type occupying the closed end of a trench (the MOTHER cell) replaced cells below, resulting in a pure genotype per trench over time (example of three trenches with randomised loading and eventual purification are shown in Fig. 5b). Thus, mixed fluorescence populations, and use of multiple specifically designed microfluidic devices (Extended Data Fig. 14 and 15c, d) allowed us to compare the effects of *metG* auxiliary expression, similarly as above, but at a single cell level.

**Fig. 5:**
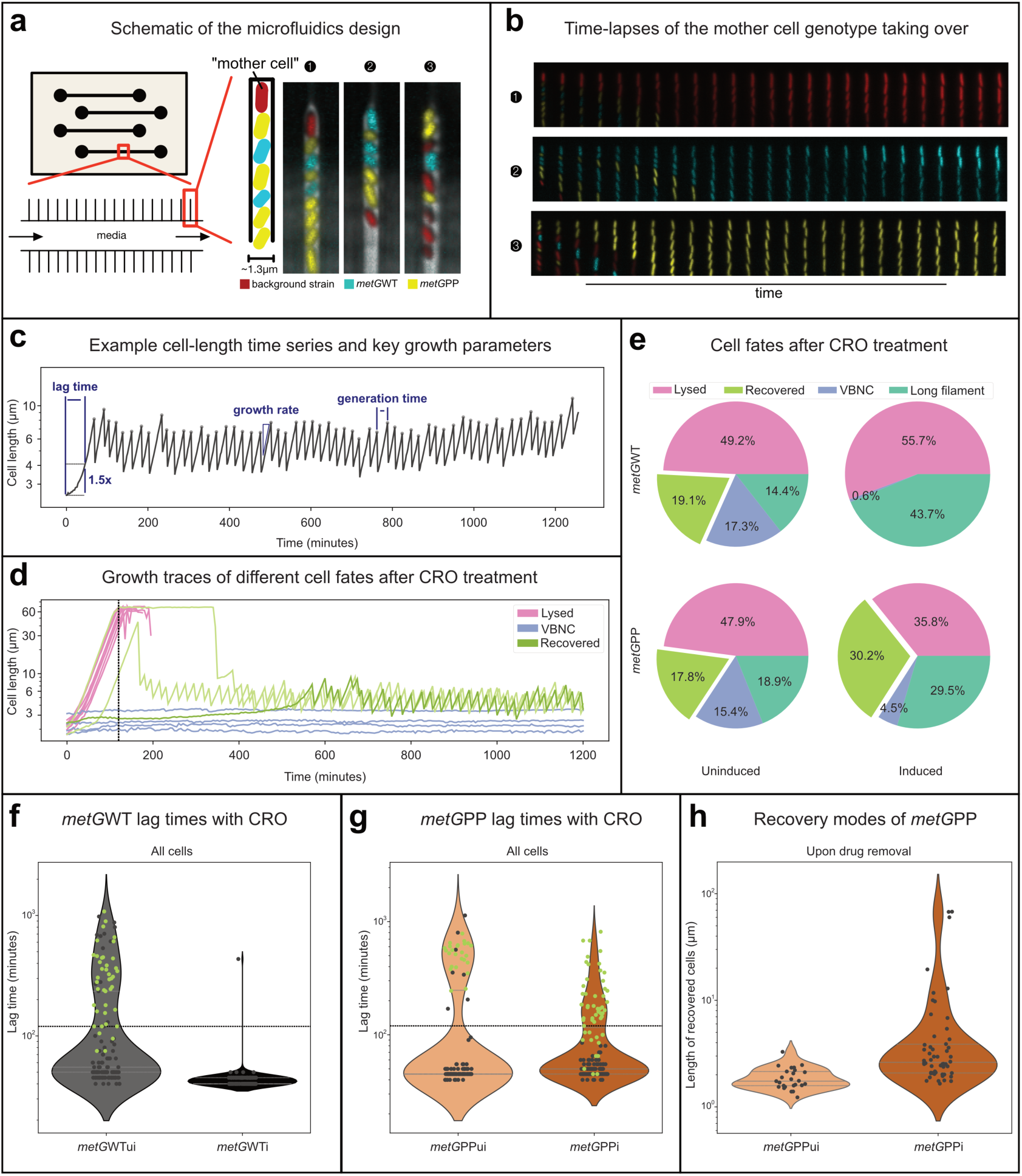
Single-cell analysis of CRO response of *metG*_WT_ and *metG*_PP_ carrying cells. **a,** A schematic of the high throughput microfluidics device for pooled single cell time lapse experiment. Individual lanes are used to load and treat cells under induced or uninduced conditions. Each lane contains narrow trenches (1.3μm wide) to house isolated cell lineages of cells from a randomised pool (*E. coli* carrying *metG*_WT_ and *metG*_PP_, and their background strain). Strains randomly occupy positions within individual trenches, but the progenies of the cell at the closed end (referred to as the mother cell) eventually occupy the trench, causing each trench to have a pure genotype. Individual cell lineages from all strains are randomly distributed across trenches and share a homogeneous treatment and growth condition from media flowing in the lane throughout specified time. Three example trench images including mixed strains at t=0, where different genotypes occupy the mother position, are shown. Each trench was identified by the “mother cell” located at the closed end of the trench. **b,** Three example timelapses of each trench shown in (**a**) are shown overtime. The trenches were occupied by the progenies of the mother cell as they were grown from t=0. **c,** Example cell-length time series and key growth parameters measured. The cell-length time series were obtained by segmentation and tracking of individual cells from stationary phase to exponential growth. From these traces, key single cell growth parameters were determined including lag time, growth rate, and generation time. **d,** Example cell-length traces leading to different fates from CRO treatment (6x MIC, 2h). A fraction of cells lyse during treatment (Pink) or form long filaments which are both considered terminal phenotypes. Blue traces are categorised as VBNC cells which show no visible signs of growth. Green traces represent cells that can recover normal growth after treatment, including cells that are dormant throughout treatment and resuscitate after the antimicrobial is removed (survivors) and cells that form short filaments and resolve into regular sized cells after the antimicrobial is removed (Short filament formers). **e,** Proportion of fates for cells after CRO treatment shifted by *metG*_WT_ and *metG*_PP_ expression. Fractions were obtained by bootstrapping approximately 200 cells for each strain. Inducing *metG*_WT_ caused most cells to lyse and reduce the fraction of VBNC (*P* < 0.001 between *metG*WTu and *metG*WTi, t-test), while inducing *metG*_PP_ led to more cells recovered from the treatment, i.e., reduced lysis and VBNC population (*P* < 0.001 between metGPPu and metGPPi, t-test). **f,** Lag time distribution of *metG*WTu and *metG*WTi cells. Recovered cells are shown as green dots with drug removal indicated by a dashed line. Most *metG*WTi cells have exited dormancy before the drug is removed. **g,** Lag time distribution of *metG*PPu and *metG*PPi cells. Recovered cells are shown as green dots with drug removal indicated by a dashed line. The *metG*PPi were able to recover cells that had exited from a wide range of time points. All the cells that exited dormancy after drug removal were able to recover. **h,** Cell size of *metG*_PP_ populations just before treatment recovery; many *metG*PPi cells resolved from filamentation through a sequence of asymmetric divisions. (For figure subsections **f,g** and **h**, the violin plots depict the distribution of data points with the horizontal lines showing the 25^th^ percentile, median, and 75^th^ percentile from bottom to the top while the edges of the violin plots show the upper and lower limits of the data points)

We then used this set up to precisely track and analyse the growth and response phenotypes of each genotype towards high-dose CRO treatment (6x MIC for 2 hours from stationary phase). An example time series of cell length from an untreated cell is shown in Fig. 5c to indicate variables at baseline in contrast with the diverse response phenotypes observed during and after CRO treatment (Fig. 5d). Diverse response phenotypes among cells after CRO treatment were classified into four distinct categories including two terminal phenotypes:

Cells that visibly lysed or 2. Cells that form long filaments; 3. Viable, But Non Culturable (VBNC), cells that did not grow during or after treatment; and 4. Recovered cells capable of normal growth after treatment. Recovered cells were further divided into two subcategories: Survivors that exited dormancy, and resumed growth, after CRO was removed and Short filament formers that formed short filaments during treatment and resolved into planktonic cells once the antimicrobial was removed through a series of asymmetric divisions. Critically, in the absence of the inducer, strains carrying both *metG*_WT_ or *metG*_PP_ plasmids showed similar CRO response profiles (see comparable phenotype proportions in Fig. 5e, Extended Data Fig. 16) highlighting their utility as controls.

The effect of expressing auxiliary *metG* was then captured through *metG*_WT_ induction which resulted in a drastic reduction of VBNCs (Fig. 5e, Extended Data Fig. 16). All cells exited dormancy and resumed growth during CRO treatment with entirely lethal outcomes (i.e. the entire population either lysed or formed long filaments, Fig. 5e). This dormancy exit was facilitated by a reduced lag time (defined as the time to reach 1.5X the dormant cell size; Fig. 5c). The comparison of lag time distributions between uninduced and induced *metG*_WT_ populations showed that expression caused all cells to exit dormancy during treatment leaving them susceptible to CRO-induced killing (Fig. 5f). This finding is consistent with prior results demonstrating that *metG* is essential for dormancy exit; an extra copy accelerates this process, while a mutated version slows it down^51,52^. Additionally, the induction of *metG*_WT_ increased the probability of dormancy exit, evidenced by the reduced fraction of VBNC cells (Extended Data Fig. 16). However, this also reduced overall survival, aligning with bulk observations (Fig. 4c).

Turning then to the effect of *metG* variation, while induction of *metG*_PP_ also reduced VBNC fractions (though to a lesser extent than induction of *metG*_WT_), it increased the fraction of recovered cells (Fig. 5e). Lag time distributions revealed that *metG*_PP_ expression led to a higher proportion of survivor cells (i.e. those with lag times longer than the treatment period) and cells that formed short filaments during treatment but resolved after CRO was removed (i.e. still recovered, but with lag times shorter than the treatment period, Fig. 5g). This distinction was further supported by cell length distributions of recovered cells immediately after CRO removal (Fig. 5h). Specifically, induction of *metG*_PP_ resulted in a greater fraction of longer cells at the point of CRO removal, which subsequently resolved into regular-sized cells, suggesting that *metG*_PP_ introduces greater variability into lag times compared to the wild-type version. This variability facilitates a bet-hedging strategy, where longer lag times allow cells to survive through treatment, and shorter lag times enable early exit from dormancy, leading to filament formation during treatment with recovery to normal division post-treatment.

### Epilogue: further real-time outbreaks underline the role of pWPMR2

During the period of this work, pWPMR2 was also involved in several new multi- and extensively drug-resistant outbreaks of *S. sonnei* that took place in high income nations, specifically, England, Spain, and Canada in 2023^52,53^. Concerningly, the Canadian outbreak was associated with shigellosis moving into a new demographic of endemic transmission, people living with homelessness, as well as an increased severity of disease^52^. The UK outbreak comprised of 120 cases within a 10-SNP-surveillance-cluster that occurred from 2022 – 2023 with most (n=116/120 isolates) being from 2023^54^). There were three isolates among this 10-SNP-surveillance-cluster in the pre-outbreak data analysed in Fig. 1 (Extended Data Fig. 17). These isolates (from 2019 and 2020) contained CRO resistance but did not contain pWPMR2. However, the 120 outbreak isolates from 2022 – 2023 contain pWPMR2 further highlighting the association of pWPMR2 in new outbreaks of highly drug-resistant bacteria. Similarly, isolates from an MSM-CRO-resistant associated outbreak in Spain in October 2023, and in 90% (n=82/91) isolates from an outbreak among people experiencing homelessness in Vancouver 2021 – 2023 also contained pWPMR2 (Extended Data Fig. 17). We also probed whether there was further evidence of pWPMR2 in lower to middle income nations by exploring data from patients who had recently travelled (an informal remote surveillance mechanism)^10,14,55^ and found 11 isolates from: Europe, Morocco, Saudi Arabia, Mexico, and Kenya which contained pWPMR2 or pWPMR2var.

## Conclusions

Here we have identified that auxiliary *metG* carried by the phage plasmid pWPMR2 is associated with evolution of bacterial lineages on an AMR trajectory; in this case *Shigella* sp. circulating in STN. Laboratory studies demonstrated that expression of auxiliary *metG* facilitated survival of antimicrobial insult and predisposed to the evolution of AMR, but at a significant cost. This is consistent with the apparent bet-hedging ecology of *metG* borne pWPMR2 in bacterial populations. Specifically, pWPMR2 is borne at low, potentially even partial copy numbers (i.e. where infecting pool would have cells with a mixture of 0 or 1 pWPMR2 copies per cell), and high variance in lag time, which would facilitate many cells avoiding the significant lag time increase, while leaving a small population able to survive antimicrobial treatment. The mutated version of *metG* borne by pWPMR2 (*metG*_PP_) not only reduced the increase in lag time, but introduced similarly stochastic population dynamics through single-cell variability in dormancy exit and cell growth under antimicrobial exposure that was not seen for the wildtype (*metG*_WT_). Thus, although conferring resistance-relevant phenotypes, pWPMR2 does not carry AMR genes, as described for some previously described phage-plasmids^56^. And finally, as a phage plasmid, pWPMR2, may not actually need its bacterial host to survive antimicrobial treatment to be passed on, with phage particles simply reinfecting new hosts once available. Collectively, this indicates that *metG*-bearing pWPMR2 has several ecological hedges to offer substantial phenotypic advantages under antimicrobial selection whilst minimising its high collateral effects.

These mechanistic insights are entirely consistent with ‘nature’s experiment’ captured in the temporospatial dynamics of pWPMR2. Specifically, pWPMR2 conferred advantages against antimicrobials that were clinically relevant for the community under study (i.e. azithromycin and ceftriaxone are used to treat many of the co-morbid STIs in MSM including gonorrhoea and chlamydia^48,49,57^). Furthermore, it’s presence preceded AMR emergence and being distributed among other clinically relevant and commensal organisms, as well as the detection of other potentially mobilisable *metG* bearing mobile genetic elements in other bacteria. Concerningly, and perhaps potentiating any services to the evolution of AMR, is that the survivor phenotype induced by auxiliary *metG*_PP_ expression might also act in other ways to advantage AMR organisms. For example, to subvert human immunological responses and contribute to chronic infections, which have been observed for some strains of sexually transmissible shigellosis^58,59^ and to subvert bacterial host defences as other phage-encoded tRNA functions have previously been described to do^60^. These alternative functions are an important avenue for future research, particularly as pWPMR2 and other tRNA-ligase encoding MGEs with the potential to contribute to AMR emergence may increasingly emerge as pathogen adaptations that persist and mobilise among microbial communities.

In the face of that possibility, there are several important avenues of future work here. Uncovering the fundamental biology of pWPMR2 and other tRNA ligase-bearing mobile genetic elements in their hosts should be explored. These studies should include aspects of transmission, evolution, and population ecology, as well as mechanistic pathways around gene regulation, expression, and functional impacts for the host. The natural distribution of genetic features that have been detected here should be explored more fully (e.g. *metG*, and other tRNA ligases on MGEs, and pWPMR2 and pWPMR2var across larger databases). For example, the screen for the distribution of pWPMR2 (Fig. 3b) did not detect pWPMR2var (Supplementary Table 6). Hence, the distribution of pWPMR2var, other *metG* (and other tRNA ligase) bearing mobile genetic elements detected (Supplementary Tables 7 and 8), alongside more targeted analyses screening datasets of other known successful AMR bacterial pathogen sublineages for copy number variation in *metG* and other tRNA ligases is warranted. These works will determine the extent to which auxiliary tRNA ligases are employed as an evolutionary adaptation in bacterial populations and provide critical information on the underpinning biological pathways.

To end, perhaps the biggest lesson from this work is the value of data driven biology for naturally evolving pathogen populations. We have, as a global public health community, sequenced >2000 pathogenic strains of bacteria containing pWPMR2 since 2004 without understanding its significance. There are two important lessons from this for future microbiological surveillance in the genomic era. The first is that we have an overly narrow focus on certain phenotypes in facing the AMR crisis. While the scientific community has known of tolerance and persistence for decades, these phenotypes are not part of routine surveillance (owing partly to a focus on patient-level outcomes). Our work highlights the need to consider more broadly what phenotypes drive bacterial lineages to epidemiological success. The second issue highlighted is an over-reliance on databases. Bacterial AMR will not stop evolving as we need to sustain life saving (though responsible) antimicrobial use. To this end, we are fortunate to now be in the genomic surveillance era facilitating enhanced taxonomic nomenclature and well-maintained feature databases. However, we also need agile systems to detect and investigate changes in pathogen genetic make-ups that we do not understand if we are to better connect pathogen evolution with public health outcomes.

## Methods

### Phylogenetic reconstruction

For this study, 3,745 *S. sonnei* isolates sampled in the UK for routine microbiological surveillance, initially sequenced as academic collaborations (isolates representative of routine surveillance from 2004 – 2014, n=329 ERR accessions) and then from routine genomic surveillance (2015 – 2020, n=3416, SRR accessions) were included (Supplementary Table 1,^14,61^). Short reads of the 3,745 *S. sonnei* isolates went through the following quality control procedures: raw read data were downloaded from the Sequence Read Archive (Bioproject: PRJNA248792, Supplementary Table 1). Reads were filtered using Trimmomatic v0.39, residual Illumina adapter sequences were removed, the first and last three base pairs in a read were trimmed, before a sliding window quality trimming of four bases with a quality threshold of 20 were applied, and reads of less than 36 bp (after trimming) were removed (ILLUMINACLIP:PE_All.fasta:2:30:10:2:keepBothReads SLIDINGWINDOW:4:20 LEADING:3 TRAILING:3 MINLEN:36)^62^. The coverage of the resulting file was estimated against the size of complete reference genome *S. sonnei* 53G (Accession: NC_016822.1) and down sampled to ∼100 x coverage. Musket v1.1^63^ was applied for k-mer spectrum read correction using a k-mer size of 31bp. The processed reads were then mapped to *S. sonnei* reference genome 53G using snippy v4.6.0 (https://github. com/tseemann/snippy), resulting in a core genome alignment of 4,988,504bp in size. After filtering out polymorphic sites within recombined regions using Gubbins^64^ with marginal ancestral state reconstruction implemented, 32,294 polymorphic sites unaffected by recombination were kept for the final phylogenetic tree reconstruction. A maximum likelihood phylogenetic tree reconstructed was performed using iqtree v2.3.6 with a General Time Reversible (GTR) substitution model with 4 gamma rate categories and 10,000 ultrafast bootstraps to provide approximately unbiased branch support values^65^. A second phylogenetic reconstruction of the evolutionary relationships among 172 representative isolates from the 3745 *S. sonnei* isolates and 123 isolates from MSM-associated *bla*_CTX-M-15_ outbreak^54^ short reads of 295 isolates went through quality control procedures as described in *Determining depth of coverage and variant calling* below. Then, a phylogenetic tree inferred similarly to above from 5,942bp polymorphic sites unaffected by recombination.

### Identification of sexual transmission network lineages

Out of 3745 *S. sonnei* isolates, 1383 isolates with demographic data combination of male, adult and without travel history to high-risk countries were given presumptive MSM status (pMSM). Ancestral state reconstruction of each internal node in the phylogenetic tree based on the pMSM status of each isolate (tree tips) was performed using MrBayes^66^ through R implementation, provided by the package MBASR^67^. The pMSM status of each isolate was refined by the reconstructed ancestral status of their most adjacent nodes, as STN lineages are typically identified by an over-representation of isolates derived from pMSM (see references below). Through plotting the frequency distribution histogram of pMSM probabilities of the most adjacent node for each isolate, pMSM probabilities greater than 0.8 were defined as pMSM (Extended Data Fig. 4). For defining STN lineages, all isolates descending from each internal node were defined as belonging to a lineage with the node being the most recent common ancestor. Then, iterating through each node in the tree, a STN lineage was defined when more than 75% of the isolates in the lineage were pMSM. This approach is an extension of, and in line with, previous work and validated surveillance practices^24^. To avoid complete reconstruction of the tree as pMSM, and reflecting the recent emergence of sexually transmissible shigellosis, only the first and second nodes that were most adjacent to the tree tips were used in STN lineages definition.

### Bacterial genome wide association analysis

Unitig-based bGWAS was run on 3,745 *S. sonnei* isolates with the 1,028 isolates belonging to MSMA lineages being a binary phenotype. Unitigs counting was performed using unitig-caller (https://github.com/bacpop/unitig-caller). The presence-absence matrix of the unitigs was associated with the binary phenotype belonging to MSMA lineages through computing terminal association scores in *treeWAS* v1.1^24^. Unitigs with the highest absolute association scores showed presence-absence patterns that were the most significantly associated with the phenotype. A minor allele frequency filter of 0.05 was applied and the unitigs with greater than 0.56 *treeWAS* terminal and subsequent association score (informed by frequency distribution histogram) were examined for further follow up.

Terminal score evaluates how a trait is distributed across the tips (leaves) of a phylogenetic tree. For binary traits, it works by tallying the four possible combinations of presence or absence of both the phenotype and the genotype among the samples. Whereas subsequent score aims at identifying associations that persist in a broad or scattered pattern across the phylogenetic tree^67^. Annotations of these unitigs were identified by mapping them to a concatenated reference consisted of the chromosome and pKSR100 plasmid of *S. sonnei* strain SRR5005407 using bwa mem v0.7.17-r1188^68^. Unitigs belonging to azithromycin resistance genes *mphA* and *ermB* were annotated using BLASTn v2.14.1+ with minimum coverage and percentage identity of 70% and 95% respectively. Mapping of the unitigs to the nine Nanopore sequenced contigs of the same reference strain was performed the same way using bwa as described above.

### Determining depth of coverage and variant calling

Illumina reads for 3745 *S. sonnei* isolates were trimmed using fastp v0.23.4^69^, using the following options, in the order as appear: auto-detection for adapter for pair-end data (-- detect_adapter_for_pe), move a sliding window of size 4bp from front (5’) to tail, drop the bases in the window if its mean quality is less than 20, stop otherwise (--cut_front), move a sliding window of size 4bp from tail (3’) to front, drop the bases in the window if its mean quality is less than 20, stop otherwise (--cut_tail), global trimming at front and tail for 15 bp (-- trim_front1 15, --trim_front2 15), detect and trim polyG in read tails in minimum length of 10 bases (--trim_poly_g), detect and trim polyX in read tails in minimum length of 10 bases (-- trim_poly_x).

The relative depth of pWPMR2 coverage was determined by mapping trimmed short reads using Bowtie2^70^ to a concatenated reference that contained the following components: SRR5005407 chromosome (Nanopore contig 1, 4 819 329bp), pKSR100 (contig 8, 65 728bp) and P-P (contig 9, 108 107bp), separated by strings of 500xN. Final destination of any read with multiple equally good alignments was randomly reported as default setting. The relative depth of coverage for each isolate was obtained from the average read depth for the pWPMR2, normalised by that of the chromosome using Samtools v 1.19^71^. PCR duplicates were marked using Samtools v 1.19^71^.

For copy number determination and variant calling in *metG,* trimmed short reads were mapped using GEM3^72^ to SRR5005407 chromosome (contig 1) only, followed by marking PCR duplicates. For *metG* variant calling, the number of reads supporting each allele in each variant position in the *metG* gene region were extracted using bcftools v1.19 mpileup command. They were then converted into .bcf file using bcftools call command, with the following options applied: output all alternate alleles in the alignments regardless of if they appear in any of the final genotype (-A), haploid (--ploidy 1), and alternative model for multiallelic and rare-variant calling (-m). For each variant position in each sample, the presence of an alternative allele was only defined when it was supported by at least 40 reads. Alternative allele proportion was calculated by the number of reads supporting the alternative allele divided by the total read depth of the position in a sample. Copy number of *metG* was obtained based on the same mapping, by computing its average read depth for each isolate, normalised by that of chromosome using Samtools v1.19. PCR duplicates were marked using Samtools^71^.

### Genetic analyses of pWPMR2

The circular topology of pWPMR2 (Nanopore contig 10) and pWMPR2var (Nanopore contig 8) was confirmed by assembling the Nanopore reads of SRR5005407 and SRR13013630 respectively using Flye v2.9.5-b1801^73^. Assembled contigs were visualised using Bandage v0.9.0.^74^. Prior assembly, adapter sequences were trimmed and reads with internal adapter were discarded using Porechop v0.2.4 (https://github.com/ rrwick/Porechop). Quality score and length of remaining reads were visualised using NanoPlot v1.43.0^75^, then reads of less than 150bp and with quality score less than 10 were filtered out using NanoFilt v2.8.0^76^.

Detection of phage sequences, plasmid specific sequences, AMR genes and defence systems were performed by running pWPMR2 through: PHASTEST v3.0^77^ using default parameters, and phage region information was obtained from detail.txt and summary.txt output files; PlasmidFinder v2.1^78,79^ with Enterobacteriales database, a minimum percentage identity of 95% and minimum coverage of 60%; and AMRfinderPlus v3.12.8^80^, DefenseFinder and AntiDefense Finder using default settings^81,82^ We determined the taxonomy of pWPMR2 using the ViPTree server^83^ to generate an all-against-all tBLASTx similarity proteomic tree against 1,512 related dsDNA prokaryotic viruses from the Virus-Host DB, which utilises complete genomes from NCBI RefSeq and GenBank^84^, also including examples from each of the five defined phage-plasmid groups within the SSU5 community^37^. For the detailed SSU5 supercommunity phylogeny, Panaroo v1.5.2^85^ was used to analyse the pangenomes (annotated using Bakta v1.8.2^86^) of the relatives detected in the NCBI nt database, as well as selected sequences identified in the literature^37^. An alignment of 39,267bp, representing 43 filtered shell genes present in 95% of sequences, was passed to IQTree v3.0.0^65^ to generate a maximum likelihood phylogenetic tree using the GTR+F+G4 model and 10,000 ultrafast bootstraps. To further explore its association with clinically relevant organisms we downloaded the genomes of its closest relatives, as found through a BLASTn search against the NCBI nucleotide collection (nt) database, selecting sequences with >95% coverage and >98% identity for direct comparison.

Five *E. albertii* plasmids were chosen for comparison with pWPMR2 as they represented the top non-*Shigella* and non-*E. coli* hits in the online nBLAST database showing highest percentage identity. To screen 66,929 genomes of enteric bacteria available from clinical cases in the UK for evidence of SSU5-like, pSLy3-like and pWPMR2-like phage plasmids and additional *metG*, a mapping approach as described previously^42^ was used. Briefly, target genes including *metG* (CP104415.1:54597..56630), *repB* (CP104415.1:22858..24072), and *repA* (NC_018843.1:59428..60492) were screened in clinical enteric bacteria derived from UK reference laboratory routine genomic surveillance, available under BioProjects PRJNA315192 (*E. coli* and *Shigella* spp.) and PRJNA248792 (*Salmonella* spp.)^42,83^.

Available data on patient travel was linked with isolates containing pWPMR2 and revealed 11 isolates (n=4 pWPMR2, n=7 pWPMR2var) with confirmed recent travel history among the surveillance data set. The relevant destinations were Belgium, Hungary, and two isolates from Spain for pWPMR2, and one from each of Morocco, Saudi Arabia, Mexico, Kenya, Bosnia and Herzegovina, and ‘Africa’ for pWPMR2var.

Structural variants between STN-lineage associated pWPMR2 and pWPMR2var, comparison of near-identical relatives of pWPMR2 in public databases, pWPMR2 and *E. albertii* plasmid comparison as well as was visualised using EasyFig genome comparison visualiser^87^ or Artemis and ACT^88^. The pWPMR2 sequence was additionally screened against ∼2M bacterial genomes in the AllTheBacteria database v0.3^89^ using LexicMap tool v0.4^90^, with a 70% coverage and 70% identity threshold. Metadata of associated hits were then retrieved using Entrez Direct (https://www.ncbi.nlm.nih.gov/books/NBK25501/). Genotype AMR analysis was done using sonneityper Genotypes (see Table 1) and the earliest year of identification of either pWPMR2 or AZM, CRO, and CIP AMR determinants was identified. Subtracting the year of AMR acquisition from the year of pWPMR2 acquisition was then used to identify the relevant time interval.

### Long read sequencing of selected isolates

Isolates were grown in TSB overnight with 200 rpm shaking at 37_°_C and 500µl of the overnight culture was then used in DNA extraction using either MasterPure total DNA/RNA extraction kit (Biosearch Technologies) or FireMonkey DNA extraction kit (Revolugen) following manufacturers’ recommendations. Extracted DNA was then quantified using a Qubit before sending off to be sequenced, analysed and annotated by commercially available Plasmidsaurus service (https://www.plasmidsaurus.com/) using Oxford Nanopore Technologies (ONT) platform.

### Analysis of metG introductions

For constructing the maximum likelihood phylogenetic tree that explored introductions of *metG*, reads from *S. sonnei* isolates containing pWPMR2 or pWPMR2var (n=650) went through quality control as described in “*Determining depth of coverage and variant calling*” above. Then were mapped to the (full length) chromosome of SRR5005407, followed by variant calling performed as for copy number determination and variant calling in *metG* (see below). Consensus sequences of *metG* in each sample were obtained using bcftools v1.19^71^ consensus command. Gene *metG* gene sequences from 285 *metG* containing plasmids from various bacterial species from PLSDB (Supplementary Data 4) were then extracted. Five plasmids with duplicate metG sequences were removed (Accessions: CP016905.1, CP022791.1, CP088831.1, NZ_CP093325.1, NZ_CP115947.1) to streamline analysis. For facilitating alignment, *metG* sequences from the 650 *S. sonnei* surveillance isolates were deduplicated using seqkit rmdup command^91^, resulting in 388 unique *metG* sequences. Together with the 285 *metG* sequences from PLSDB, 673 *metG* sequences were aligned using Kalign3 v3.4.0^92^, followed by phylogenetic reconstruction using iqtree v3.0.1 with parameters as described as above^65^. The number of variant sites from the alignment was obtained using SNP-sites v 2.5.1^93^.

### Analysis of plasmid and bacteriophage databases

PLSDB database version 2023_11_23 was downloaded from https://ccb-microbe.cs.uni-saarland.de/plsdb/plasmids/download/ and annotated using Bakta version 1.8.2^86^ with the ‘full’ database. A list of COG categories for tRNA synthetases was downloaded from the COG database and used as search terms in the Bakta output .gff files. To avoid potentially misclassified chromosomal sequences in PLSDB, sequences encoding ≥3 tRNA synthetases were removed from analyses of tRNA synthetase-encoding plasmids. Plasmid types were extracted from PLSDB metadata. Analyses were conducted in R using tidyverse^94^. This analysis was repeated with a curated bacterial prophage database: prokaryotic dsDNA virus sequences downloaded from Virus-Host DB^83^ (n=5631). In addition, the annotations provided with the INPHARED phage database^95^ (n=28665 excluding RefSeq genomes, downloaded from https://millardlab.org/2025/03/06/phage-genomes-march-2025/) were searched for the presence of tRNA synthetase/ligase products (Supplementary Table 8).

### Construction of E. coli strains

*metG*_WT_ sequence obtained from the chromosomal copy of *metG* from *S. sonnei* SRR5005407 and the *metG*_PP_ sequences obtained from the pWPMR2 of *S. sonnei* SRR5005407 were used to synthesise the respective versions of *metG* using GeneArt custom synthesis platform (ThermoScientific). The synthesised genes were then cloned into the inducible pTrcHis2A vector using the same service and the two resulting plasmids were then transformed into *E. coli* SB8 (a *motA* deletion variant of *E. coli* MG1655^96^) and selected using the ampicillin marker (100µg/ml) on the vector. Transformed *E. coli* strains were used in the downstream experiments with 1mM IPTG (Millipore Sigma, UK) as the inducer to induce the expression of the *metG* genes.

### Sub-MIC growth curves

Minimum Inhibitory Concentrations (MIC) for each of the *E. coli* SB8 strains carrying the two different versions of *metG* genes on pTrcHis2A vector were determined using E-strips (Liofilchem, Italy) and/or broth dilution method for azithromycin (0.25µg/ml), ciprofloxacin (0.0078125 µg/ml) and ceftriaxone (0.0625µg/ml). Those strains were then grown in biological and technical triplicates in TSB containing half of the MIC concentrations of each of the three antimicrobials with and without induction of the *metG* variants. The OD_600nm_ values were recorded every 15 minutes using a BioTek Synergy H1 multi-mode plate reader. The resulting data were then analysed using gcplyr R package^97^ or the shiny-based version of miLAG^98^ and graphs were generated using ggplot2 R package^99^.

### Experimental evolution study

*E. coli* SB8 containing *metG*_WT_ and *metG*_PP_ on a pTrcHis2a plasmid backbone and the parent strain without any plasmids were streaked out on TSA plates containing 100µg/ml ampicillin for the plasmid carrying strains and without antimicrobials for the parent strain and incubated overnight at 37_°_C to obtain isolated colonies of each of the *metG* variant carrying *E. coli* and the parent strain. These isolated colonies were then inoculated into individual wells of 96 well plates containing either no antimicrobials or AZM, CIP or CRO at a concentration of ½ MIC value for each and treated as an individual population. Each 96 well plate contained 47 wells with TSB with IPTG at a final concentration of 1mM and a control well, 47 wells with no IPTG and a control well. These 96 well plates were then incubated at 37_°_C with shaking at 200 rpm for 24 hours before transferring 1% (v/v) into a fresh 96 well plate containing twice the concentration of each of the antimicrobials from the overnight plate. This was carried on for 11 days with the final concentration of each antimicrobial reaching 512x MIC. At the end of each of the 24-hour incubation periods, each of the wells were scored for growth using the OD_600nm_ values obtained by end point reads from a BioTek Synergy H1 plate reader and the number of populations survived was plotted against the antimicrobial concentration using the ggplot2 R package. Statistical analysis for the survival of each of the populations was performed using the survival R package (https://CRAN.R-project.org/package=survival).

### Microfluidic single-cell experiment

The microfluidic devices were fabricated following the protocol described in Wedd et al^51^. On the day before the experiment, the bacterial strains (*metG*_WT_, *metG*_PP_, and background) were inoculated into TSB and incubated overnight with 200 rpm shaking at 37_°_C for 18 hours. After incubation, the cultures were pooled in equal proportions and loaded into the device. Individual strains were identified based on their chromosomally integrated fluorescence protein markers, which produced bright signals in dedicated fluorescence channels. Just before the image acquisition, the two syringes with flow medium (with and without inducer) were prepared, as well as with and without CRO (6x MIC, 0.4µg/ml). These media were connected to designated flow lanes in the microfluidic device and switched during the experiment. Time-lapse multi-colour imaging was conducted to monitor the exit dynamics and post-exit growth and division of the cells. Images were captured using a Nikon Eclipse Ti2 inverted microscope with a Hamamatsu C14440-20UP camera. All images were saved in Nikon’s ND2 file format and processed to extract single-cell growth traces using the pipeline described by Wedd et al^51^, which uses a combination of synthetic training images and machine-learning models for single-cell segmentation^100^. Cells were tracked across frames using a custom script (https://github.com/erezli/MMLineageTracking). The extracted time-series data were analysed to determine key growth parameters, such as lag time, using custom Python scripts.

## Acknowledgements

This work was funded by BBSRC (BB/V009184/1) and MRC (MR/X000648/1) project grants. JPJH is supported by an MRC Career Development Award (MR/W02666X/1). The research in the Bakshi lab was supported by the Wellcome Trust Award (grant number RG89305), a University Startup Award for Lectureship in Synthetic Biology (grant number NKXY ISSF3/46), an EPSRC New Investigator Award (EP/W032813/1) and a seed fund from the School of Technology at University of Cambridge. CJ, CRB, KSB, LCEM and PR are affiliated to the National Institute for Health Research Health Protection Research Unit (NIHR HPRU) in Gastrointestinal Infections at University of Liverpool in partnership with the United Kingdom Health Security Agency (UKHSA), in collaboration with University of Warwick. SN and PR are affiliated with the NIHR HPRU in Genomics and Enabling Data at University of Warwick in partnership with the UKHSA, in collaboration with University of Cambridge and University of Oxford. The views expressed are those of the author(s) and not necessarily those of the NHS, the NIHR, the Department of Health and Social Care or UKHSA. PR acknowledges the Research/Scientific Computing teams at The James Hutton Institute and NIAB for providing computational resources and technical support for the UK’s Crop Diversity Bioinformatics HPC (BBSRC grant BB/S019669/1), use of which has contributed to the results reported within this article. The authors would like to thank Professor Wei Shen for his generous support regarding installing and running AllTheBacteria database and Lexicmap tool. For the purpose of open access, the authors have applied a Creative Commons Attribution (CC BY) licence to any Author Accepted Manuscript version arising from this submission.

## Data availability statement

Nucleotide sequences of pWPMR2 and pWPMR2var obtained using Nanopore sequencing are deposited with accession numbers PQ180110 and PQ521219 (respectively) in GenBank. All metadata for the strains and their individual genome accession numbers are provided in Supplementary Table 1.

## Unique Biological material availability statement

Requests to access unique biological materials used in this manuscript will be considered and should be directed to the corresponding author.

## Code availability statement

Unique code used to track cells across frames in the microfluidic single cell experiments is available in GitHub (https://github.com/erezli/MMLineageTracking) and the custom python scripts used to extract time-series data such as lag time are available upon request from the corresponding author.

## Author contributions

Conceptualization – KSB, JPJH, SB, RL

Data curation – KSB, LC, SCB, CRB, CJ, RL

Formal analysis – CRB, YLT, JPJH, LS, GBB, MDS, CEC, SCB, PR, SB, RL

Funding acquisition – KSB, JPJH, CJ, MDS, LC, SB

Investigation – KSB, LC, YLT, JPJH, LS, SN, MDS, CEC, SCB, PR, SB, RL

Methodology – KSB, LC, YLT, PR, SB. RL

Project administration – KSB, MDS

Resources – KSB, CJ

Supervision – KSB, LC, JPJH, SCB, CJ, SB

Visualisation – KSB, CRB, YLT, GBB, MDS, CEC, LS, JPJH, SB, RL

Writing – original draft – KSB, YLT, JPJH, GBB, MDS, CEC, SB, RL

Writing – review & editing – KSB, LCEM, LC, CRB, YLT, JPJH, SN, MDS, CEC, SCB, PR, CJ, SB, RL, GBB, LS

